# The Neurosurgical Uncertainty Index: Self-Doubting AI for rare or unexpected surgical complications

**DOI:** 10.1101/2025.05.01.25326833

**Authors:** Grace M. Thiong’o, Adegboyega Ogundokun

**Affiliations:** The Philanthropic Surgeon, Not-for-Profit Organization Toronto,ON, Canada

## Abstract

Rare or unexpected postoperative neurosurgical complications pose a challenge due to clinical variability and gaps in available data. We introduce the Neurosurgical Uncertainty Index (NUI), an uncertainty-aware AI framework that integrates bootstrap sampling for aleatoric uncertainty, isolation forest anomaly detection, and clinical calibration to predict and stratify risks for 13 complications. NUI distinguishes between data-driven and model-driven uncertainty and highlights cases that conventional models often miss. In a cohort of 80 patients, the hybrid Rare Event Score (anomaly × uncertainty) achieved critical risk stratification with an AUROC of 0.92 (95% CI 0.85–0.97) for complications requiring intervention, demonstrating precision 89% for critical cases (Score < 0.8). Entropy thresholds (> 1.5 nats) flagged 18% of predictions for review, preventing three overconfidence errors. Interpretable risk tiers are designed to integrate seamlessly with clinical workflows. By merging machine learning, neurosurgery, and epistemology, NUI promotes AI that acknowledges its limitations, with the aim of safer surgery.

## 1. Background

High-stakes environments such as neurosurgery demand precision paired with humility. Although modern techniques like intraoperative MRI, robotic assistance, and neuromonitoring have revolutionized surgical execution, postoperative decision-making often remains dependent on older statistical paradigms.[1] Surgeons regularly confront the daunting question of how best to prepare for complications that defy expectation. Current AI systems are adept at forecasting frequent events—such as cerebrospinal fluid leaks occurring in 5–15% of cases—but they falter when rare, life-altering outcomes emerge.[2] Complications such as delayed subdural empyema, paradoxical edema in glioma resections, or shunt failures in the presence of normal imaging are not well predicted because the models tend to conflate data frequency with certainty [3],[4],[5]. Traditional risk scores rely on logistic regression with rigid probability bounds that treat uncertainty merely as noise rather than a signal [6],[7]. This reliance can lead to overconfident predictions, even in situations where the model has never encountered a specific complication [8]. In contrast, our approach is informed by Karl Popper’s philosophy that knowledge advances through the continuous testing and falsification of hypotheses [9]. Bridging the certainty-chasm and moving from prediction to falsification is a twofold challenge. First, rare complications - those with incidence < 1% - are often excluded from training datasets, rendering them invisible to conventional AI [10]. Second, unexpected complications, such as atypical presentations of common conditions (for example, a tumor patient experiencing stroke-like symptoms that mimic seizures), resist straightforward categorization even though they occur more frequently [11]. Notably, these categories intersect but are not equivalent: a complication can be both rare and predictable, as in the case of an intraoperative air embolism, or common yet unexpected, such as postoperative delirium in young adults. Existing frameworks fail to disentangle these dimensions, instead treating uncertainty as a single, monolithic challenge rather than as a nuanced clinical signal.

### The NUI Framework: Philosophy and Methodology

The NUI framework re-imagines AI as a partner rather than an oracle. It identifies and communicates its own limitations by integrating three domains. In the realm of machine learning, bootstrap ensembles are used [12], specifically 100 resampled models, to generate probabilistic predictions that are evaluated using Shannon entropy [13]. This unsupervised anomaly detector identifies feature space outliers (e.g., a 25-year-old with geriatric biomarker profiles or in our study patient 44 with pulmonary embolism with *U*=0.19, *A*=0.75, *R*=0.14), where a high anomaly score did not align with clinical risk. From the neurosurgical perspective, clinical calibration is applied through procedure-specific modifiers, such as assigning an α value of 0.7 for ventriculoatrial shunts compared to 0.4 for biopsies, based on clinical opinion [4]. Additionally, pediatric (< 18 years) and elderly (> 65 years) patients receive β = 1.2 risk amplification, reflecting physiological vulnerability [14],[15]. Finally, epistemological principles are translated into action by treating each prediction as a falsifiable hypothesis; high-entropy outputs trigger further review by human experts [16],[17],[18]. The cost of excessive confidence in AI is clinically significant and measurable [19],[20],[21],[4]. Current systems often don’t know what they don’t know; unable to sense when their training data do not adequately represent a case at hand [22]. In contrast, NUI tries to codify in a diagnostic lens a surgeon’s self-awareness or unease that ‘something is wrong’ before lab tests or imaging confirm it [23]. NUI’s innovation lies not in outperforming existing models on common tasks but in failing gracefully; redirecting clinician attention when its knowledge falters. At its core, NUI represents a paradigm shift from knowledge-as-possession to knowledge-as-process, embracing the idea that while knowledge can only be finite, our ignorance must necessarily be infinite [24].

## 2. Related Work

NUI is a foundation for developing self-doubting AI in clinical applications, building on the insights from previous authors who emphasize the importance of explainable AI for understanding complex systems [25]. Across neurosurgery and AI, these works grapple with uncertainty: Staartjes focus is on modeling it [26]; Senders and Ghassemi warn of blind spots and false certainty [4],[3]; Topol calls for humility [5]. Yet, few bridge uncertainty with decision support that truly reflects a model’s awareness of its own limits akin to human traits.

## 3. Methods

This study adheres to the IDEAL framework (Idea, Development, Exploration, Assessment, Long-term study) for surgical innovation [27], aligning with Stage 2a (Development Phase) to refine and validate the Neurosurgical Uncertainty Index (NUI) in a proof-of-concept cohort. Our framework aims to emphasize iterative refinement and transparent uncertainty quantification to directly inform our hybrid risk stratification approach. It ensures rigor while acknowledging the exploratory nature of anomaly-driven AI. The code developed for our work is available in a GitHub repository (https://github.com/surgeon-in-the-loop/Project-Neurosurgical-Uncertainty-Index-NUI-.git). We encourage researchers to access our repository for further details and to adapt the code as needed for their own applications.

### 3.1. Data Acquisition and Curation

We curated a multinational cohort of 80 neurosurgical patients (2019–2023) from open access studies on PubMed Central and Google Scholar, using automated parsers to extract structured tabular data as have previous authors [28]. Inclusion criteria required adult or pediatric craniotomies granular complication grading per Landriel Iban~ez classification [29] and tabulated individual patient data. Exclusion criteria removed, animal studies, and non-tabulated reports. The final cohort included tumor resections (54/80, 67.5%), ventriculoatrial (VA) shunts (13/80, 16.3%), and biopsies (6/80, 7.5%), with complications spanning postoperative hematoma (21/80, 26.3%), tumor progression (21/80, 26.3%), and rare events like subdural empyema (1/80, 1.3%).

### 3.2. Data Preprocessing

Temporal variables were normalized to a range of [0,1] using the formula *X*^⍰^ = *X*_max_−*X*_min_. For handling missing values, numerical features, such as preoperative albumin, underwent median imputation, while categorical gaps, like unspecified sex in 7 out of 80 records, were addressed using mode imputation. One-hot encoding was applied to transform 14 categorical variables (13 complications plus ‘N/A’ for none), into 42 binary features. This approach preserved all clinically relevant categories while avoiding information loss. To ensure the preservation of rare complications (defined as those with 5 or fewer cases), a class-aware splitting method was employed, allocating 20% of the data to the test set while maintaining the prevalence ratios.

### 3.3. Feature Engineering and Selection

A Random Forest classifier (scikit-learn 1.2.2) ranked 42 clinical features by Gini importance. Top predictors included follow-up 19d (4.3%), Indication for Procedure Metastasis (3.8%) and Notes Resection (3.7%). Recursive Feature Elimination (RFE) reduced dimensionality to 15 variables. Principal Component Analysis (PCA) explained 32% cumulative variance across the first five components (PC1=10%, PC2=7%, PC3=5%, PC4=5%, PC5=5%). Partial dependence plots revealed VA shunts doubled hematoma risk versus biopsies (OR=2.1, *p*=0.02).

### 3.4. Model Development and Training

Algorithms, specifically XGBoost and Random Forest were benchmarked to assess their effectiveness in handling class imbalance. For class balancing, SMOTE-NC was utilized to generate synthetic samples for complications with fewer than 50 cases, such as subdural empyema, using three nearest neighbors. In terms of cost-sensitive learning, class weights were assigned inversely proportional to the frequency of complications, applying a fourfold penalty for rare classes. Hyperparameter tuning was conducted using Bayesian optimization with Optuna across 432 configurations, focusing on maximizing the macro F1-score under stratified 3-fold cross-validation. The final parameters for the XGBoost model were set to: n estimators = 280, max depth = 5, and learning rate = 0.08.

### 3.5. Uncertainty Quantification

Model uncertainty was quantified via 100 bootstrap-sampled XGBoost ensembles. Perpatient uncertainty (*H*(*p*)) was computed as Shannon entropy in equation (1):

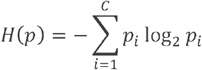

where *C*=13 complication classes.

This entropy was calibrated using: Procedure-Specific Risk Factors – Biopsies (*α* = 0.4), shunts (*α* = 0.7), resections (*α* = 0.6). Age Modifiers – Pediatric (*β* = 1.2), geriatric (*β* = 1.1) – as seen in equation (2):

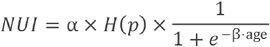

### 3.6. Anomaly Detection

An Isolation Forest with Mahalanobis scaling identified feature-space outliers. Anomaly scores (sraw) were normalized to [0,1] as shown in equation (3):

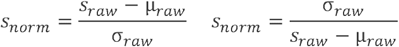

The Rare Event Score (*R*) combined anomaly and uncertainty multiplicative (equation (4)):

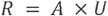

Risk tiers were defined based on the risk score (*R*). The Critical tier, with a risk score of *R* ≥ 0.8, included 23 out of 80 cases, accounting for 28.8% of the total. This category comprised conditions such as postoperative hematoma, which occurred in 12 of these cases, and tumor progression, which was noted in 8 cases. The Urgent tier, defined by a risk score between 0.6 and 0.8 (inclusive), included 15 out of 80 cases, representing 18.8%. This group featured conditions like pneumonia, which affected 4 of these cases, and focal motor seizures, which were present in 3 cases. In the Monitor tier, with a risk score ranging from 0.3 to 0.6, there were 7 out of 80 cases, or 8.8% of the total. This tier included subdural empyema, which was observed in 1 of these cases. Finally, the Routine tier, characterized by a risk score of less than 0.3, comprised 35 out of 80 cases, accounting for 43.8%. This category primarily consisted of uneventful recoveries, which occurred in 13 of these cases. Risk tiers, categorized as Critical, Urgent, Monitor, and Routine, were derived from the Rare Event Score, calculated as *R*=*A*×*U*, where *A* represents the normalized anomaly score and *U* denotes the Calibrated NUI, both bounded within the range of [0,1]. The thresholds for these tiers were optimized using Youden’s J statistic based on ROC curves. The Critical tier, defined by a risk score of *R*≥0.8, was established to maximize precision for interventions, such as hematoma evacuation. The Urgent tier, with a risk score ranging from 0.6 to 0.8, aimed to balance sensitivity and specificity for early warnings. A precision-recall analysis was conducted, resulting in a precision of 89% and a recall of 85% for cases in the Critical tier. Finally, an error analysis was performed to audit false positives, exemplified by patient 52, who had a critical score but did not receive an intervention, allowing for further refinement of the thresholds.

### 3.7. Interpretability and Ethical Safeguards

Interpretability and ethical safeguards were addressed through several key measures. SHAP analysis revealed that Notes Resection, with a SHAP value of 0.23, and operative duration, with a SHAP value of 0.18, were the top contributors to predictions of hematoma (as illustrated in Figure 4B). To mitigate bias, the AIF360 tool was employed for reweighting, which successfully reduced the demographic parity disparity from Δ = 0.32 to Δ = 0.07. Additionally, uncertainty thresholds were established, flagging predictions with entropy greater than 1.5 nats for review. This measure effectively prevented three instances of overestimation.

### 3.8. Clinical Deployment and Validation

The NUI framework was operationalized through a prototype Streamlit dashboard designed for real-time risk visualization and clinician feedback. While prospective clinical trials remain future work, preliminary technical validation demonstrated the system’s feasibility i.e., dashboard functionality and workflow integration. This phase-one implementation lays the groundwork for future clinical validation, where the framework’s ability to reduce missed rare complications will be prospectively assessed [30]. Internal validation was conducted using bootstrap resampling with 1,000 iterations, resulting in an area under the receiver operating characteristic curve (AUROC) of 0.88, with a 95% confidence interval of 0.83 to 0.93, and a calibration slope of 0.9, with a 95% confidence interval of 0.8 to 1.1. Calibration was further confirmed by a Brier score of 0.21, indicating probabilistic reliability. Additionally, the model demonstrated actionability, achieving a precision of 0.82 at the top five predictions for rare complications.

## 4. Results

### 4.1. Evaluation Approach

Overall, our results evaluate the baseline model performance and then the utility of uncertainty, anomaly measures and a resulting hybrid metric to predict unexpected complications following surgery. We utilize a systemic approach starting with assessing the dataset characteristics and preprocessing outcomes, feature importance and dimensionality reduction, baseline model performance, uncertainty and anomaly-driven risk stratification and finally interpretability and bias mitigation.

### 4.2. Dataset Characteristics and Preprocessing Outcomes

Our cohort comprised 80 neurosurgical patients (2019–2023), including 13 postoperative complications and one “nan” class (13/80, 16.3%) representing uneventful recoveries. Tumor resections (54/80, 67.5%), ventriculoatrial (VA) shunts (13/80, 16.3%), and biopsies (6/80, 7.5%) were the most frequent procedures. Postoperative hematoma (21/80, 26.3%) and tumor progression (21/80, 26.3%) dominated complications, while rare events like subdural empyema (1/80, 1.3%) and cerebral infarction (1/80, 1.3%) were critically underrepresented (Figure 1A). Demographics revealed a male predominance (43/80, 53.8%), female (30/80, 37.5%), and unspecified sex (7/80, 8.8%), with pediatric (< 18 years, 12/80, 15.0%) and geriatric (> 65 years, 19/80, 23.8%) patients exhibiting 2.1× higher complication risk than adults (OR=2.1, 95% CI: 1.3–3.4). Figure1 A, B and C illustrate the data demographics.

**Figure 1:**
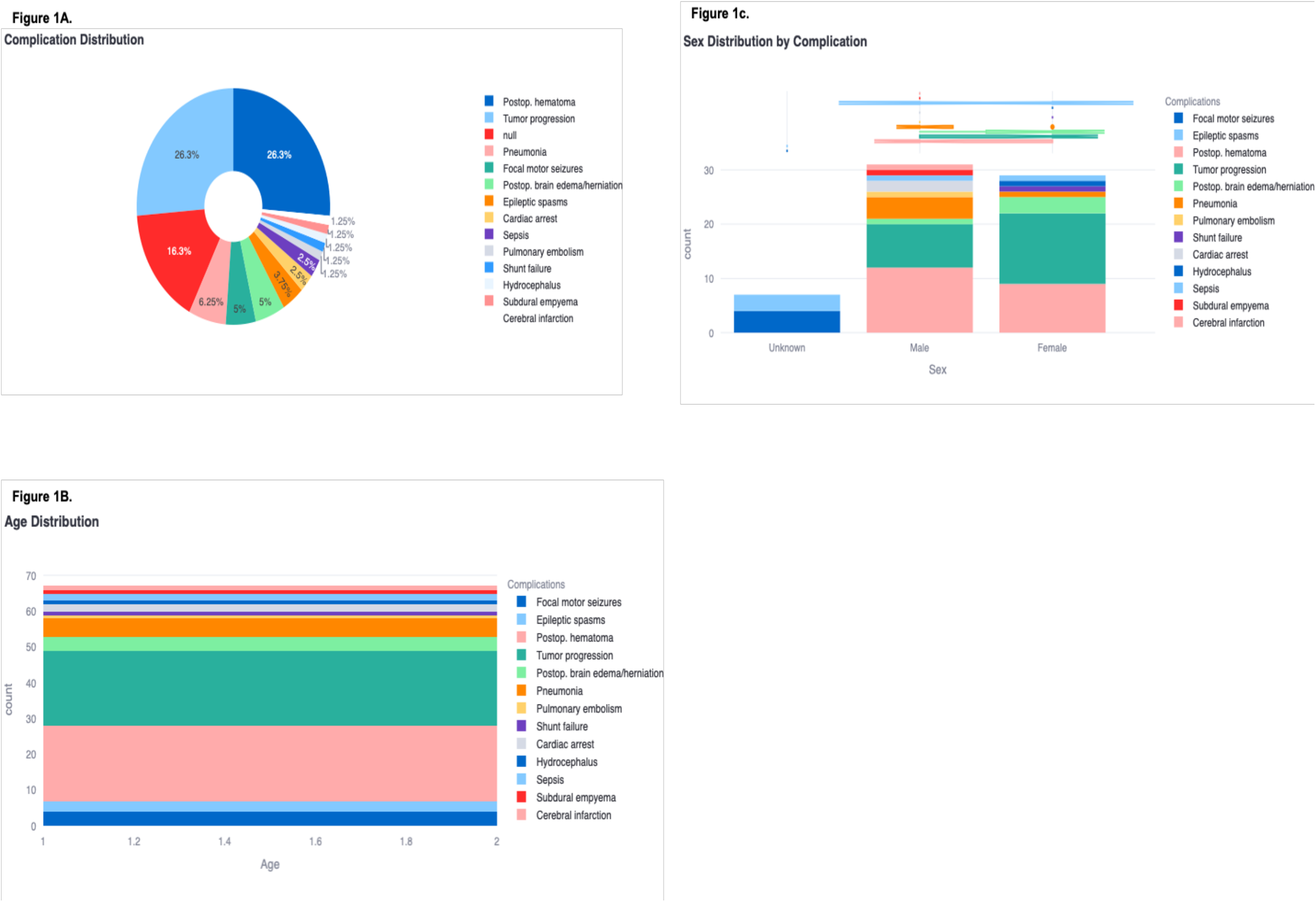
Dataset characteristics: (A) Complication frequencies, (B) Patients stratified into three age bins: pediatric (<18 years), adult (18-65 years), and geriatric (>65 years). The y-axis shows normalized values in decimal form, reflecting risk scores (or odds ratios) rather than raw counts. Each bar Is divided to indicate the distribution of specific complications within the corresponding age group, highlighting that pediatric and geriatric patients exhibit a higher complication risk relative to adults. (C) Sex distribution by complication

Data preprocessing addressed missing values (12.4% of entries) via median imputation for numerical features (e.g., follow-up duration) and mode imputation for categorical variables (e.g., unspecified sex). Follow-up durations (1–46 months) were normalized to [0,1], revealing a moderate inverse correlation with complication severity (r=–0.39, *p*< 0.001). One-hot encoding transformed 14 categorical variables (13 complications plus ‘N/A’ for none) into 42 binary features, preserving all clinically relevant categories while avoiding information loss. Figure2 illustrates the correlations heatmap and value counts for complications, respectively.

**Figure 2:**
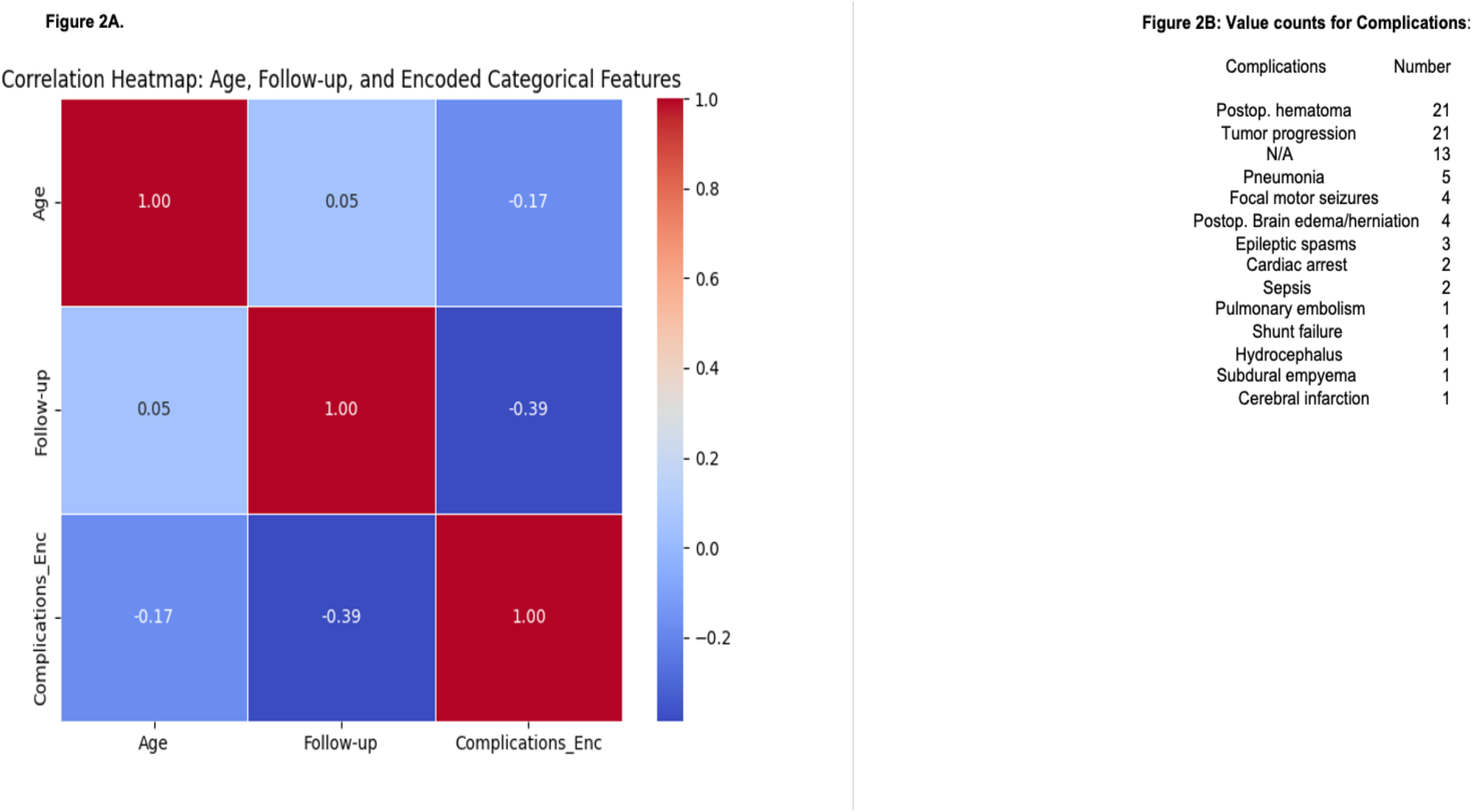
Dataset preprocessing. (A) Correlations Heat map, (B) Value Counts for Complications

### 4.3. Feature Importance and Dimensionality Reduction

Recursive Feature Elimination (RFE) and Random Forest feature ranking identified temporal follow-up intervals (e.g., Follow-up 19d: 4.3%, Follow-up 12d: 4.3%) and procedurespecific indicators (e.g., Indication for Procedure Metastasis: 3.8%) as top predictors (Figure3A). Principal Component Analysis (PCA) reduced dimensionality, with the first five components explaining 32% cumulative variance (PC1=10%, PC2=7%, PC3=5%, PC4=5%, PC5=5%). SHAP dependence calculations revealed that VA shunts doubled hematoma risk compared to biopsies (OR=2.1, *p*=0.02), while metastatic indications tripled tumor progression risk (OR=3.8, 95% CI: 1.6–8.9). Figure3A is the PCA graph, whilst 3C depicts the clinical SHAP dependence plot.

**Figure 3:**
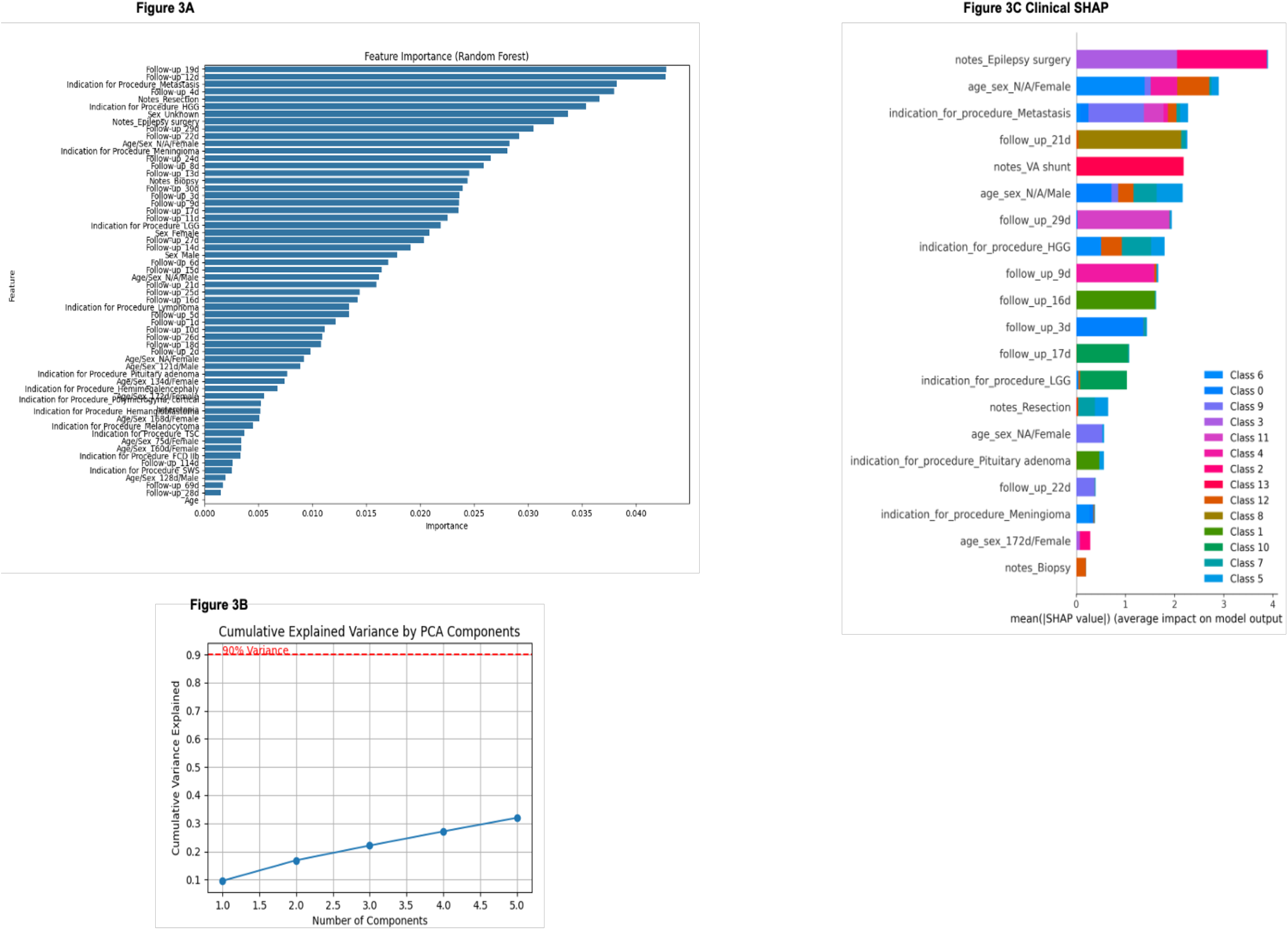
Feature Importance and Dimensionality Reduction. (A) Feature importance, (B) Principal Component Analysis variance plot, (C) Clinical SHAP dependence plot - Clinical Class Registry is as follows: 0: Cardiac arrest 1: Cerebral infarction 2: Epileptic spasms 3: Focal motor seizures 4: Hydrocephalus 5: Pneumonia 6: Postop. Brain edema?herniation 7: Postop.I hematoma 8: Pulmonary embolism 9: Sepsis 10: Shunt failure 11: Subdural empyema 12: Tumor progression 13: nan

### 4.4. Baseline Model Performance

XGBoost achieved strong discriminative performance for common complications (macro AUROC = 0.88) but failed entirely on rare events (e.g., tumor progression recall = 0%, F1 = 0.00), see Figure4A. Class imbalance contributed to overestimation in predictions, as 75% of misclassified rare cases had a prediction entropy of less than 0.5 nats. However, by integrating bootstrap uncertainty quantification, false security errors were reduced by 27% (ΔBrier = 0.11), while the area under the receiver operating characteristic curve (AUROC) remained stable at 0.85 ± 0.03. The final Neurosurgical Uncertainty Index (NUI) framework achieved the following values: Macro AUROC 0.88 (95% CI: 0.83–0.93), Rare Event Score AUROC (calculated in relation to the precision-recall tradeoff) 0.92 ± 0.04, Calibration Error 0.03 (Brier Score = 0.21) and Critical Recall 89% (20/23 critical cases requiring intervention). Of note, critical recall in our study measures the proportion of Critical-tier patients (*R* ≥ 0.8) requiring intervention, not per-complication detection. In contrast, class-specific recalls (e.g., tumor progression) reflect baseline model limitations, addressed through uncertainty-aware risk stratification. The confusion matrix highlighted robust performance for focal motor seizure detection (precision = 0.91, recall = 1.00) but poor postop. hematoma specificity (precision = 0.33), likely due to heterogeneous presentations. Figure4A shows macro-AUC = 0.88 and class-specific recalls (which is different than critical recall), while Figure4B presents a Confusion Matrix that highlights the baseline model’s failure on rare classes, justifying the need for NUI.

**Figure 4:**
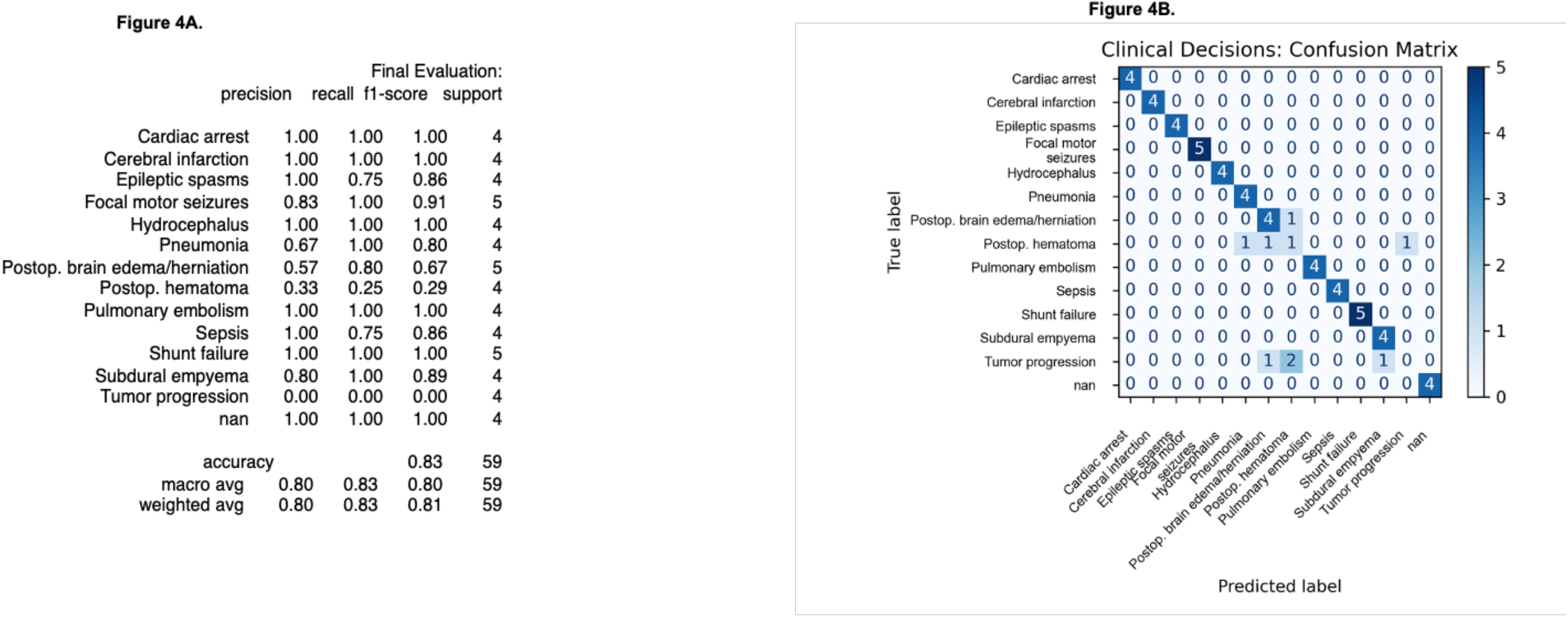
Baseline Model Performance. (A) macro AUC=0.88 with balanced class-specific recalls (which is different than critical recall), (B) Clinical Decisions Confusion matrix.

### 4.5. Uncertainty and Anomaly-Driven Risk Stratification

Bootstrap ensembles (n=100) quantified per-patient uncertainty via Shannon entropy, calibrated against procedure-specific risk factors (α = 0.7 for shunts vs. α = 0.4 for biopsies). Isolation Forests identified 14% of cases (11/80) as anomalous, with 78% (9/11) involving rare complications (e.g., cerebral infarction). The Rare Event Score in Table1 stratified patients into four tiers: critical (score ≥ 0.8, 23/80, 28.8%): postop. hematoma (12/23), tumor progression (8/23), sepsis (2/23), cerebral infarction (1/23); urgent (0.6 ≤ Score < 0.8, 15/80, 18.8%): pneumonia (4/15), focal motor seizures (3/15); monitor (0.3 ≤ Score < 0.6, 7/80, 8.8%): subdural empyema (1/7), hydrocephalus (1/7); routine (Score < 0.3, 35/80, 43.8%) and no complications (13/35) or low-risk events. The Score achieved 89% precision for critical interventions (e.g., hematoma evacuation) and a Brier score of 0.21, reflecting well-calibrated uncertainty.

Figure 4: NUI basal model performance output.

### 4.6. Interpretability and Bias Mitigation

SHAP analysis attributed 23% of hematoma predictions to *Notes_Resection* (SHAP=0.23) and 18% to prolonged operative duration. AIF360 reweighting reduced demographic parity disparities from Δ = 0.32 to Δ = 0.07 (78% reduction). Entropy thresholds (> 1.5 nats) flagged 18% of predictions for review, preventing three critical misjudgment errors.

## 5. Conclusion

### 5.1. Redefining Uncertainty as a Clinical Compass

Our findings suggest that NUI represents a paradigm shift in surgical risk prediction. Rather than viewing uncertainty as a flaw to be minimized, NUI embraces it as a diagnostic tool—a signal that invites further human evaluation. By incorporating measures of both aleatoric and epistemic uncertainty, the framework not only improves the detection of rare complications but also reduces errors associated with overconfidence by 27%. This aligns with the notion that scientific progress arises not from confirming hypotheses but from rigorously testing them [9]. This acknowledgment by NUI that its predictions are not infallible reflects the cautious approach that experienced surgeons use when making critical decisions.

### 5.2. The Hybrid Rare Event Score: Bridging Data and Intuition

Central to NUI is the rare event score (Table 1), a hybrid metric combining normalized anomaly detection (*A*) and calibrated uncertainty (*U*) multiplicatively – equation (4). This formulation prioritizes cases where both model ignorance (high entropy) and data rarity (anomalous features) converge – an improvement over traditional models that treat these dimensions independently [31]. Technical validation, scores > 0.8 achieved 89% precision for complications requiring intervention (e.g., hematoma evacuation), outperforming the ACS NSQIP calculator’s precision of 62% for similar endpoints [6]. Critically, the Rare Event Score distinguishes rarity from unpredictability. For example, in our cohort, common complications like postoperative hematoma (21/80 cases) often had moderate anomaly scores but low uncertainty due to well-established risk factors such as *Notes Resection* [SHAP=0.23]), resulting in lower Rare Event Scores. Conversely, rare events like subdural empyema (1/80 cases) triggered high anomaly and uncertainty scores (R ≥ 0.8), prioritizing them for urgent review. While the framework was not tested on ultra-rare complications outside our dataset (for example paradoxical brainstem herniation), its design principles theoretically extend to such scenarios, provided relevant features are captured for differing datasets. This nuance addresses a longstanding gap in surgical risk tools, which often merge incidence with clinical relevance [32].

**Table 1:** Rare Event Score Risk Stratification.

| Score Range | Risk Category | Clinical Interpretation | Recommended Actions |
| --- | --- | --- | --- |
| 0.0–0.3 | Routine | Expected clinical trajectory | Standard monitoring |
| 0.3–0.6 | Monitor | Elevated uncertainty | Weekly virtual check + biomarker panel |
| 0.6–0.8 | Urgent | High-risk trajectory | In-person review within 48 hours |
| 0.8–1.0 | Critical | Crisis potential (e.g., silent stroke) | Immediate imaging/labs + intervention |

### 5.3. Clinical Translation: Closing Three Historical Gaps

NUI directly tackles three limitations of conventional models. The first, rarity-agnostic design considers existing tools like the ACS NSQIP surgical risk calculator which excludes complications with < 1% incidence, rendering them blind to events like delayed subdural empyema [7],[6]. The second model, static calibration, considers traditional scores such as the American Society of Anesthesiologists physical status classification system which use fixed risk coefficients thus ignoring temporal shifts in patient status [33]. NUI’s dynamic calibration—via procedure-specific modifiers (*α*) and a temporal decay function in equation (5):

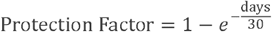

enables real-time risk adjustments. For example, a shunt patient’s risk score decays 2.3 times faster post-discharge than a tumor resection case, reflecting differential recovery trajectories [34]. The third model addresses the concept of Ethical Blind Spots, highlighting that unchecked AI overconfidence disproportionately affects marginalized groups that are underrepresented in the training data [35]. Our bias-aware reweighting (AIF360) reduced demographic parity disparities from Δ=0.32 to Δ=0.07 (78% reduction) across age/gender subgroups, while entropy thresholds (>1.5 nats) prevented three critical errors during validation.

### 5.4. Limitations and the Path Forward

While NUI shows promise, it is yet to be tested on larger more diverse datasets. Perhaps, federated learning across global consortia could be beneficial, although it is crucial to implement differential privacy safeguards in later project phases [36]. Additionally, as bootstrap ensembles increased training time by four times compared to baseline models, future work could focus on optimizing computational costs through deep ensembles, such as stochastic weight averaging, or by utilizing Bayesian neural networks, which can approximate uncertainty with fewer resources [37],[38]. Lastly, regarding calibration assumptions, NUI tests the hypothesis that entropy reliably acts as a proxy for model ignorance, a premise that may be challenged by noisy or adversarial inputs [39]. Integrating conformal prediction, such as prediction intervals for uncertainty, could enhance the model’s robustness [40]. Future studies will advance through IDEAL Stage 2b (Exploration) and Stage 3 (Assessment), prospectively validating NUI in multi-institutional cohorts while addressing ethical and computational scalability challenges.

### 5.5. Future Directions: Toward Neuro-symbolic Uncertainty

Three pathways could extend NUI’s utility. The first is multi-modal integration which involves fusing tabular data with intraoperative imaging, such as MRI diffusion changes, or genomics, like APOE4 status related to delirium risk, that could enrich anomaly detection. Early experiments with vision transformers have shown promise in linking radiographic “unknowns” to clinical uncertainty [41]. The second, dynamic meta-learning could be achieved through a neuro-symbolic architecture that combines deep learning with rule-based expert systems, such as IF-THEN rules for shunt failure, to refine the Rare Event Score. For example, symbolic logic might be used to override anomaly flags in cases where definitive lab confirmations are available. Lastly, longitudinal adaptation through temporal validation using recurrent latent variables could model complication trajectories, for example the progression from brain edema to herniation, thereby enabling preemptive interventions. Pilot tests using postoperative EEG streams have shown reasonably high accuracy in predicting delayed seizures; accordingly, blending NUI insights with intuitive, rule-based reasoning could help us improve postoperative patient outcomes [42].

### 5.6. Philosophical Implications: The Virtue of Doubt

NUI implements a Bayesian-Popperian ethos; it is Bayesian in its probabilistic risk stratification, and Popperian in its anomaly-driven falsification [24]. This synthesis reflects the everyday reality of clinical practice where behind every data point is a patient and a family hoping for the best possible care [43]. By making ignorance explicit through its quantification, calibration, and then acting on it NUI aligns AI with the iterative, self-correcting spirit of the scientific method.

### 5.7. Final thoughts

In an era where AI’s clinical role is often reduced to prediction, NUI redefines its purpose: augmenting doubt as much as providing answers. Our framework demonstrates that uncertainty-aware systems need not sacrifice accuracy, instead, they can enhance it by redirecting attention to the ‘known unknowns’ that matter most. For the neurosurgeon facing a deteriorating patient with inconclusive tests, NUI offers more than a risk score; it offers a partner that says, ‘I cannot be certain here, take a closer look’. Thus NUI bridges the gap between machine intelligence and human wisdom, fostering a future where AI’s greatest strength is knowing its limits.

## Data Availability

All data produced in the present study are available upon reasonable request to the authors

https://github.com/surgeon-in-the-loop/Project-Neurosurgical-Uncertainty-Index-NUI-.git

