## Supplementary figures and images for "The Neurosurgical Uncertainty Index: Self-Doubting AI for rare or unexpected surgical complications"

### Supplementary Material

**Supplementary Figure 1; Anomaly shap**

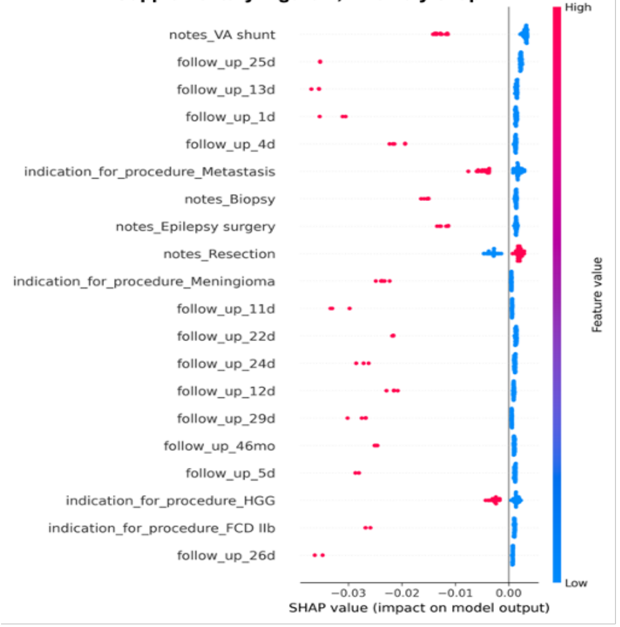


**Supplementary Figure 2**

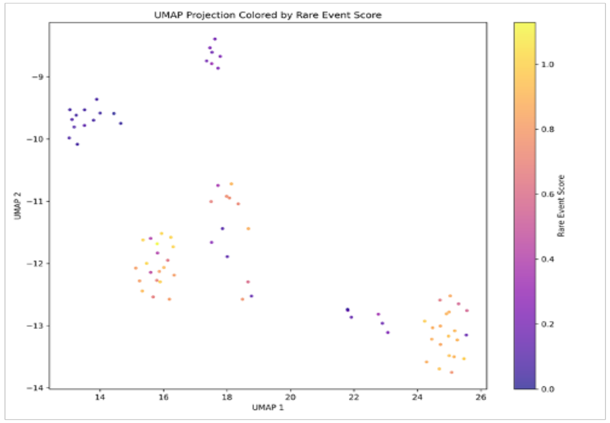


**Supplementary Figure 3:**

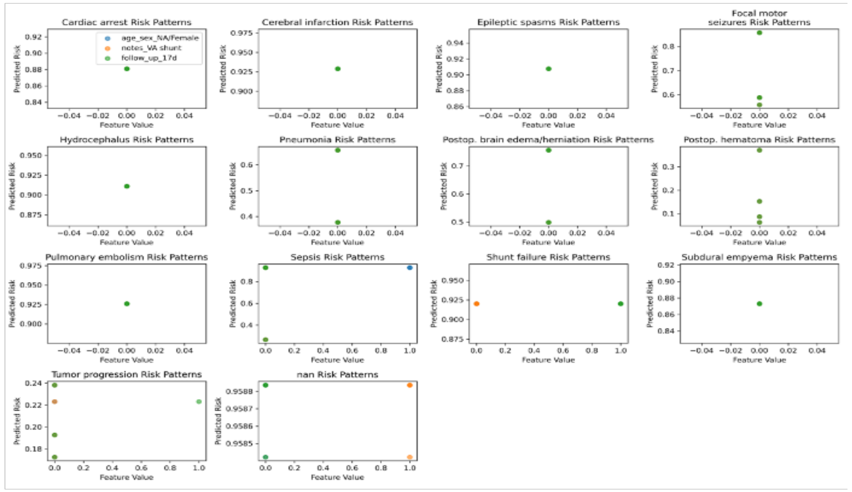
